# On the effect of age on the transmission of SARS-CoV-2 in households, schools and the community

**DOI:** 10.1101/2020.07.19.20157362

**Authors:** E Goldstein, M Lipsitch, M Cevik

## Abstract

**Background:** There is limited information on the effect of age on the transmission of SARS-CoV-2 infection in different settings, including primary, secondary and high schools, households, and the whole community. We undertook a literature review of published studies/data on detection of SARS-CoV-2 infection in contacts of COVID-19 cases, as well as serological studies, and studies of infections in the school setting to examine those issues.

**Results:** Our literature review presents evidence for significantly lower susceptibility to infection for children aged under 10 years compared to adults given the same exposure, for elevated susceptibility to infection in adults aged over 60y compared to younger/middle aged adults, and for the risk of SARS-CoV-2 infection associated with sleeping close to an infected individual. Published serological studies also suggest that younger adults (particularly those aged under 35y) often have high cumulative rates of SARS-CoV-2 infection in the community. Additionally, there is some evidence of robust spread of SARS-CoV-2 in secondary/high schools, and there appears to be more limited spread in primary schools. Some countries with relatively large class sizes in primary schools (e.g.Chile and Israel) reported sizeable outbreaks in some of those schools, though routes of transmission of infection to both students and staff are not clear from current reports.

**Conclusions:** Opening secondary/high schools is likely to contribute to the spread of SARS-CoV-2, and, if implemented, it should require both lower levels of community transmission and greater safeguards to reduce transmission. Compared to secondary/high schools, opening primary schools and daycare facilities may have a more limited effect on the spread of SARS-CoV-2 in the community, particularly under smaller class sizes and in the presence of mitigation measures. Efforts to avoid crowding in the classroom and other mitigation measures should be implemented, to the extent possible, when opening primary schools. Efforts should be undertaken to diminish the mixing in younger adults to mitigate the spread of the epidemic in the whole community.

## Introduction

Among those infected with SARS-CoV-2, elderly patients have had the most severe outcomes, including the highest death rates, whereas infected younger persons, particularly children aged 1-18y, if symptomatic at all, are far more often mildly ill *(1,2)*. While this age-dependent pattern of illness severity has become well-established, the roles of different age groups in transmission has not been as clear. Recently, evidence has accumulated that susceptibility to infection generally increases with age, e.g. *(3,4)*. This, however, does not suggest that the oldest individuals necessarily play the leading role in the spread of SARS-CoV-2 in the community – in fact, serological studies suggest that younger adults, particularly those aged under 35y often experience the highest cumulative rates of infection *(5-9)*, possibly due to age-related differences in mixing. Additionally, there is uncertainty as to the role of different age subgroups of children in the spread of SARS-CoV-2, including how susceptibility to infection varies in different age groups of children, and how it compares to susceptibility to infection in different age groups of adults. The effect of the ongoing and future openings of schools and higher-educational institutions on the spread of infection requires a better characterization of transmission dynamics in different age groups. Here, we review the relevant evidence based on household, school and community studies, and draw some conclusions regarding the relevant public health policies.

### Age variation in susceptibility to infection given contact

We undertook a literature review using the Living Evidence on COVID-19, a database collecting COVID-19 related published articles from Pubmed and EMBASE and preprints from medRxiv and bioRxiv, with MESH terms including (“child” OR “age” OR “aged”) AND (“contacts” OR “household” OR “transmission” OR “susceptibility” or “contact tracing”) to assess the susceptibility to and transmission in different age groups *(10)*. We included nine studies where estimates of either secondary attack rate, susceptibility to, or odds ratio for infection in different age groups were present, and where the setting for the contact, if varying among contacts (e.g. household vs. other) was adjusted for (as a covariate in a model) in those estimates – the latter was done to reduce the effects of heterogeneity in exposure on those estimates.

#### 1. There is evidence that susceptibility to infection in children under the age of 10y is significantly lower compared to adults

In this subsection we present the included studies that assess relative susceptibility for children vs. adults, describe the potential biases in those studies, and present a way to circumvent those biases to estimate susceptibility in children aged under 10y vs. adults.

##### Studies of SARS-CoV-2 infection in close contacts

Several studies found much lower secondary attack rates (measured by PCR-positive cases among contacts) in children – using different age cutoffs of children up to age 20y -- compared to adults (Table 1). In a hospital-based study near Wuhan, China *(11)*, the household secondary attack rate in children under the age of 18y was 4%, compared with 17.1% for adults, OR= 0.18 (95% CI (0.06,0.54)). In a Guangzhou, China study *(3)*, the multivariable odds ratio for infection in persons aged under 20y vs. persons aged over 60y was OR=0.23(0.11,0.46)), while for persons aged 20-59y vs. persons aged over 60y it was OR=0.64 (0.43,0.97). In another hospital-based study in Wuhan, China *(12)*, the secondary attack rate in tested adult household contacts was 58.6% vs. 11.2% in pediatric household contacts (age unspecified), OR= 11.21 (2.54,102.9). For household contacts of confirmed cases in Zhuhai, China *(13)*, the secondary attack rate in persons aged under 19y was 16.1%, while in persons aged 19-60y it was 37% (OR=0.33(0.09,1)), whereas in persons aged over 60y, the secondary attack rate was 41.9% (OR=1.23(0.48,3.08) relative to persons aged 19-60y). In a study of household contacts in Bnei Brak, Israel *(14)*, the secondary attack rate was 58.3% in adults aged 18-47y, 32.5% in children aged 5-17y, OR=0.35 (0.12,0.97) relative to adults, and 11.8% in children aged 0-4y, OR=0.09 (0.01,0.49) relative to adults. In a Chinese study *(4)*, the attack rate among contacts of confirmed cases (not only household) was lower in children under the age of 15y compared to adults aged 15-64y (multivariable OR= 0.34 (0.24 to 0.49)). A study modeling transmission within households in Israel*(15)* found that susceptibility in children under the age of 20y was 0.45(0.40, 0.55) that of adults. Additionally, in the same study, data on household secondary attack rates in different age groups (Figure 4 in *(15)*) suggests that secondary attack rates in children aged 0-4y and 5-9y were significantly lower compared to children aged 10-14y, and less than half that for children aged 15-19y.On the other hand, in a smaller study from Shenzhen, China *(16)*, the household secondary attack rates in children aged <18y was 35.3%, while in adults aged 18-49y it was 27.3%, and in persons aged over 50y was 20.7%. The differences between age groups (OR=0.69 (0.17,2.99) for adults aged 18-49y vs. children, and OR=0.49 (0.10,2.28) for persons aged over 50y vs. children) failed to reach statistical significance. Finally, the multivariable analysis of 1286 close contacts of 391 index cases in Shenzhen, China *(17)* suggests that infection rates in close contacts were similar across age groups (except for higher point estimates in the older age groups). However, 298/391 of index cases in this study were travelers, with joint travel being associated with an odds ratio of 7.1 (1.4,34.9) for infection in close contacts. This suggests rather irregular patterns of transmission, such as the possibility of acquisition of infection outside the household (including at the source of travel) for non-index cases, etc., making the interpretation of the age-specific susceptibility differences in *(17)* difficult.

**Table 1:** Odds ratios and relative susceptibility to infection in contacts in different age groups for studies *(3,4,11-17)*. H: household contacts; C: community contacts; All: all contacts were tested; S: symptomatic contacts were tested; U: unspecified (ref. 16). Also see the caveats in the *Potential biases and susceptibility for children under 10y* subsection.

| Study | Age groups |  |  |
| --- | --- | --- | --- |
| (4)<br>*H+C, All* | <15y<br>OR=0.34 (0.24, 0.49) | 15-64y<br>OR=1 (ref.) | Over 65y<br>OR=1.67 (1.12,1.92) |
| (3)<br>*H+C, All* | <20y<br>OR=0.23 (0.11,0.46) | 20-59y<br>OR=0.64 (0.43,0.97) | Over 60y<br>OR=1 (ref.) |
| (13)<br>*H, All* | <19y<br>OR=0.33 (0.09,1) | 19-60y<br>OR=1 (ref.) | Over 60y<br>OR=1.23 (0.48,3.08) |
| (16)<br>*H, U* | <18y<br>OR=1 (ref.) | 18-49y<br>OR=0.69 (0.17,2.99) | Over 50y<br>OR=0.49 (0.10,2.28) |
| (14)<br>*H, All* | 0-4y<br>OR=0.09 (0.01,0.49) | 5-17y<br>OR=0.35 (0.12,0.97) | Over 18y<br>OR=1 (ref.) |
| (11)<br>*H, All* | <18y<br>OR= 0.18 (0.06,0.54) |  | Over 18y<br>OR=1 (ref.) |
| (15)<br>*H, All* | <20y<br>Susceptibility = 0.45 (0.40,0.55) |  | Over 20y<br>Susceptibility=1 (ref.) |
| (12)<br>*H, S* | Children<br>OR=1 (ref.) |  | Adults<br>OR=11.21 (2.54,102.9) |
| (17)<br>*H, All* | No age-related differences in susceptibility were found |  |  |

##### Potential biases for the estimates of susceptibility in children vs. adults

Besides the potential bias for study *(17)* described above, estimates of susceptibility in children vs. adults based on household attack rates may be downwardly biased because certain adult-adult contacts may be more sustained than adult-child contacts: that is, higher secondary attack rates in adults vs. children may reflect greater exposure in addition to differences in susceptibility given the same exposure. A serological study of a SARS-CoV-2 outbreak on a US Navy ship *(18)* found that cumulative incidence of infection was higher among persons who reported sharing the same sleeping berth with a crew member who had positive test results (65.6%), compared with those who did not (36.4%), OR = 3.3 (1.8,6.1). This suggests that among contacts of an index case in a household, the spouse might face an additional risk for infection due to shared bed. In fact, for adult contacts in the Wuhan study *(11)*, the attack rate among spouses of index cases was 27.8% (25/90), whereas the secondary attack rate among other adults in the household was 17.3% (35/202), with the relative risk (RR) for infection for a spouse of the index case vs. a non-spouse adult household contact being RR=1.60(1.02,2.51). In the Zhuhai study *(13)*, 12/23 spouses of index cases were infected, compared to 31/89 of other adult contacts of index cases, with the relative risk for infection being RR=1.50(0.92,2.43). We note that the vast majority of detected COVID-19 index cases in household studies are adults, with a sizeable share (depending on the study, e.g. *(11,13)*) of adult household contacts being the spouses of the indices.

A second source of bias in the estimated susceptibility of children vs. adults is the possibility of the following scenario: a child is infected first in a household and transmits to an adult, but because the child has no or few symptoms, the child is not tested initially, and the adult is considered the household index. Then, if the original (actual household index) infection in the child is not subsequently detected by RT-PCR, this would bias the secondary attack rate (SAR) in pediatric contacts downward. On the other hand, if that original infection in the child is subsequently detected, it would be classified as a secondary infection in a study, biasing the SAR in children upward.A third source of bias, likely downward, in the estimated susceptibility of children vs. adults is related to detection of secondary cases. It is known that duration of SARS-CoV-2 shedding increases with age in adults *(19-21)*; children may have shorter duration of viral shedding in respiratory samples compared to adults, potentially leading to a lower likelihood of detection of infection by RT-PCR compared to adults.

##### Estimation of the relative susceptibility to SARS-CoV-2 infection in children aged under 10y vs. adults

An approach that avoids these potential biases is to compare infection in different age subgroups of children among household contacts. For one study we reviewed, the data are available to compare secondary attack rates in children aged 0-4y and 5-9y to those of older children; SAR in these younger groups are less than half those children aged 15-19y in *(15)*. Given that there is no evidence that 15-19y olds are more susceptible than adults, this within-childhood comparison can support the inference that children aged under 10y are at most half as susceptible as adults. Additionally, for household studies *(11,13)*, the secondary attack rate in non-spouse adult household contacts of index cases is more than twice as high as the secondary attack rates in pediatric household contacts. This, together with evidence of elevated susceptibility in older age groups of children compared to younger ones *(15)* further supports the concept that with equivalent exposure, the risk of infection in children aged under 10y is at most ½ that of adults.

#### 2. There is evidence that susceptibility to infection in adults aged over 60y is elevated compared to younger/middle-age adults

One study *(3)* estimates that for household contacts among persons aged 20-59y, the OR for infection is 0.64 (0.43,0.97) compared to household contacts aged over 60y. Another study *(4)* estimates that for household contacts among persons aged over 65y, the OR for infection is 1.67 (1.12,1.92) compared to household contacts aged 15-64y. A smaller study *(13)* estimates the OR for infection in persons aged over 60y vs. 19-60y as 1.23 (0.48,3.08). Another smaller study *(16)* estimates the OR for infection in persons aged over 50y vs. 19-49y as 0.70 (0.17,2.62).

Table 1 summarizes the above estimates for the relative susceptibilities/odds ratio for infection in different age groups. Note that in addition to the biases described in the *Potential biases and susceptibility for children under 10y* subsection, a some of the studies tested only symptomatic contacts, which can further bias the relative susceptibility for adults vs. children upward as infections in children are less likely to be symptomatic. Fortunately, all but two studies reviewed tested both symptomatic and asymptomatic contacts. One *(12)* tested only symptomatic contacts and may suffer this bias. Testing criteria in study *(16)* couldn’t be determined from the description.

### Age variation in infectivity

There is limited evidence in the literature regarding age-related differences in infectivity. Dattner et al. *(15)* estimated the relative infectivity for children aged under 20y compared to adults as 0.85 (0.65,1.1). Data from the 54 households in the Netherlands *(22)* suggest lower point estimates for transmissibility of infection to close contacts from children aged under 19y, and higher point estimates for adults aged over 70y compared to persons aged 19-69y. A large study of contacts (household and other) of COVID-19 cases in South Korea suggests that for adult index cases, household secondary attack rates generally increase with the age of the index *(23)*. For index cases aged 0-9y, the household secondary attack rate is lower than for adult indices. For index cases aged 10-19y, the household secondary attack rate is significantly higher than for index cases aged 20-29y, 30-39y, 40-49y; however, persons aged 10-19y reported by far the smallest number of contacts suggesting that some of the index cases aged 10-19y were potentially infected in the household rather than being index cases *(23)*.The notion that infectivity generally increases with age is also supported by studies of viral shedding. Evaluation of viral shedding in adult patients hospitalized with COVID-19 in Wuhan, China and Zhejiang, China *(19-21)* shows that older age was a factor associated with prolonged virus shedding time of SARS-CoV-2.

#### Potential biases in infectivity studies

As in studies of age-specific infectiousness, there may be errors in ascertaining the direction of transmission, leading to confusing differences in infectiousness with differences in susceptibility (if the direction of transmission is erroneous).

### Age variation in seroprevalence

We reviewed all seroprevalence studies in the Living Evidence on COVID-19 database *(10)* (as well as ref. *(5)*) that provided age-specific seropositivity rate estimates. Twelve studies were included in the analyses, though some of them, e.g. studies of blood donors *(5,6,24)* may suffer from selection biases.Several serological studies estimate that younger adults, particularly those aged under 35y have the highest seroprevalence of all or nearly all age groups. Serological studies in different regions in England, Rio de Janeiro, Brazil, Tokyo, Japan, as well as Heinsberg, Germany estimate that SARS-CoV-2 seroprevalence is highest in adults aged under 35y *(5-8)*. A serological study in a slum community in Buenos Aires, Argentina found the highest seroprevalence to be in adolescents aged 14-19y *(25)*. In Belgium, persons aged 20-30 years had the highest estimated seroprevalence in persons aged below 80 *(24)* (with the oldest individuals likely affected by outbreaks in long-term care facilities). In Geneva Switzerland, persons aged 20-49 years had the highest estimated seroprevalence, followed closely by those aged 10-19 years *(9)*.

Serosurveillance of adults outside grocery stores in New York State, USA found that rates of infection in individuals aged over 55y were significantly lower than in persons aged 18-54y *(26)*; the highest infection rates in New York State were in persons aged 45-54y. A serological study in Spain used a point-of-care test, and an immunoassay, each having sensitivity below 90% *(27)*. For the immunoassay, the highest seroprevalence was estimated in persons aged 20-49y, whereas for the point-of-care test, the highest seroprevalence was in persons aged over 50y *(27)*. We also note the high prevalence of multigenerational families in Spain that likely affected the relative incidence of infection in older adults. A serological study in Iceland has the highest estimate for the cumulative incidence of infection in persons aged 40-50y (Figure 2E in *(28)*), though a sizeable proportion of individuals with serologically confirmed infection were travelers, and rates of seropositivity in different age groups in this study were low. A study of seroprevalence in ten US locations *(29)* found that the age group with the highest seroprevalence estimate varies by location, with highest rates observed in adults aged 19-49y in three locations, adults aged 50-64y in three locations, adults aged 65+y in two locations, and those aged 0-18y in two locations. A serological study in Los Angeles County found similar seroprevalence estimates in different age groups of adults *(30)*.

#### Potential biases in seroprevalence studies

Sources of sera for seroprevalence surveys are almost always unrepresentative of the source population, to an extent. In particular, when individuals are asked to participate in the study in different venues for obtaining specimen (grocery store parking lots, blood donors), those who consent may differ from those who decline. We also note that serosurveys cumulate the exposure over a particular time period, and many serosurveys included time periods during which schools and workplaces were closed, potentially reducing by varying amounts the exposure of different age groups from what would be “normal”.

## Discussion

In this review, we examine evidence in the literature and provide a comprehensive overview regarding the role that age plays in the transmission of infection in different settings. Below, we review some public health issues related to our findings.

A study in a town in northern France found high rates of seroprevalence for anti-SARS-CoV-2 antibodies among high school students, and an even higher seroprevalence among the school staff following an outbreak in that school *(31)*. A study in Jerusalem, Israel found high rates of PCR-detected SARS-CoV-2 infection in students in grades 7-9 as well as in staff during a school outbreak *(32)*. A serological study in Santiago, Chile following an outbreak that led to a school closure found high rates of anti-SARS-CoV-2 antibody seroprevalence in both the staff, as well as secondary school students (significantly higher than among high school students in that school) *(33)*. This, combined with evidence of relatively high seroprevalence for anti-SARS-CoV-2 antibodies in persons aged 10-19y *(9,25)*, and evidence of elevated susceptibility to infection in children aged 10-19y compared to children aged 0-9y *(15*) suggests caution in opening secondary/high schools as it may lead to substantial rates of infection in the school setting and potentially have an effect on the spread of SARS-CoV-2 in the community.

We found evidence for a significantly lower susceptibility to infection in children under the age of 10y compared to adults and older adolescents. This, combined with evidence of a more limited spread of SARS-CoV-2 in primary schools compared to high schools *(34,31)*, as well as evidence of limited spread in smaller studies of close contacts of younger children, e.g. *(35,36)* suggests the possibility of a more limited effect of opening primary schools on SARS-CoV-2 transmission in the community compared to opening secondary/high schools. We note that classroom crowding and other factors related to social distancing in classrooms/schools may play a role in the spread of SARS-CoV-2 in primary schools. For example, Denmark, Austria, and Finland have various mitigation strategies implemented as part of opening of primary schools *(37)*; moreover, those countries have notably smaller classroom sizes in primary schools compared to England, Israel and Chile *(38)*.Therefore, the spread of SARS-CoV-2 in primary schools in former countries may be more limited (media publication *(39)*) compared to Chile, Israel and England. Primary schools saw a limited opening in England on June 1 (beginning of week 23), with a gradual increase in school outbreaks reported on weeks 23, week 24, and weeks 25-28 *(40)*. Following the opening of primary schools in Leicester, England, the percent of tests positive for SARS-CoV-2 saw the greatest increase in children aged under 19y (pp. 21-22 in *(41)*), suggesting a relative increase in SARS-CoV-2 incidence in children compared to the overall community. Israel experienced a marked growth in SARS-CoV-2 incidence in June, 2020, with elevated rates of COVID-19 in children compared to adults (media publication *(42)*) and sizeable outbreaks reported in some elementary schools *(39)*. A serological study *(33)* was performed to analyze an outbreak in a large private school in Santiago, Chile that has 14 grades (from pre-school to high school) and large class sizes (25-27 students in pre-school, 36-38 students in the rest of the school). While some of the infections recorded in this study *(33)* could have taken place after the school was closed on March 13, the fact that seroprevalence in pre-school and primary school students is higher than in high school students in *(33)* combined with evidence of higher susceptibility to infection in older children compared to younger ones *(15)* suggests infections in younger students before the school closure. We should note that there is uncertainty as to the routes of transmission in outbreaks in primary schools in England, Israel and Chile (e.g. the possibility that some pre-school students were infected by their caregivers or in the household in Chile *(33)*), though the size of the outbreaks, particularly in Israel and Chile points to the school setting as the venue for transmission.

All of this suggests that opening of primary schools should be accompanied by other measures including reducing crowding and otherwise mitigating transmission *(37,39)*. To facilitate the openings of pre-schools and primary schools, more work is needed to better characterize the spread of SARS-CoV-2 in pre-schools and primary schools, including the effect of class size and the feasibility/effectiveness of social distancing measures, sources of infection in students and staff, and the spread of infection from those institutions to households and other settings. Indeed, school re-openings, like other policies, should be undertaken with a knowledge that the policy may need to change as data and experience accumulate.

Elevated susceptibility to infection in individuals over 60y of age *(3,4)* given same exposure, and infectivity no lower than in younger persons *(22,23)* may contribute to the spread of infection in older adults. Some examples of this are Spain *(27)*, which has a high prevalence of multigenerational families that likely affected the relative incidence of infection in older adults, and New York City *(26)*, where seroprevalence in persons aged over 55y (21.5% (19.6%–23.5%)) was on par with the seroprevalence in persons aged 18-34y (21.8% (19.2%–24.4%)). Another example is the Italian town of Vo. That town has a fairly old age distribution (Table 2 in *(43)*, with 85.9% of residents sampled), and the highest incidence of infection during both population samplings was observed in older persons *(43)*. Further effort is needed to better understand how SARS-CoV-2 infection spreads in the elderly population, and how to better protect the elderly persons living outside long-term care facilities including facilitation of diminished social interaction (such as allocating certain time slots for grocery shopping among the elderly only, etc.).

A number of serological studies, e.g. *(5-9)* have the highest seroprevalence estimates belonging to younger adults (particularly those aged under 35y), as well as adolescents aged 14-19y *(25)*. While this finding is not uniform across all the published serological studies, and some of these are based on blood donor studies with potential selection bias, the ongoing relaxation of social distancing measures, and the high prevalence of younger adults and older adolescents among reported COVID-19 cases in different countries and regions in the northern Hemisphere during the summer of 2020 (e.g. *(44,32)*) suggest that efforts should be made to reduce the number and intensity of contacts in those age groups in order to mitigate the spread of the epidemic in the whole community, particularly during the Fall of 2020. Further work is needed to better understand the spread of SARS-CoV-2 in younger adults and older adolescents, including the settings and circumstances when active spread of infection takes place, the types of mitigation efforts that can be undertaken to limit the spread of infection in younger adults and older adolescents, and the effect that the above infections have on the epidemic dynamics in the whole community.

## Conclusions

We found evidence that compared to younger/middle aged adults, children aged under 10y have significantly lower susceptibility to SARS-CoV-2 infection, while adults aged over 60y have elevated susceptibility to infection, and they merit extra efforts for protection against infection.There is evidence of robust SARS-CoV-2 spread in schools for older children, and caution should be exercised in opening middle/high schools, particularly in terms of requiring both lower levels of community transmission and greater safeguards to reduce transmission. Compared to secondary/high schools, opening of primary schools and daycare facilities may have a more limited effect on the spread of SARS-CoV-2 in the community, particularly under smaller class sizes and in the presence of mitigation measures *(37)*. Efforts to avoid crowding in the classroom and other mitigation measures should be implemented, to the extent possible, when opening primary schools. Finally, given the high seroprevalence for SARS-CoV-2 in younger adults (particularly those aged under 35y) in a number of serological studies, as well as elevated rates of COVID-19 in younger adults in a number of locations during the summer of 2020, efforts should be undertaken to diminish the mixing in younger adults to mitigate the spread of the epidemic in the whole community.

## Data Availability

This paper uses publicly available data that can be accessed through the following URLs:
http://ww11.doh.state.fl.us/comm/_partners/covid19_report_archive/state_reports_latest.pdf
https://assets.publishing.service.gov.uk/government/uploads/system/uploads/attachment_data/file/901803/Weekly_COVID19_Surveillance_Report_week_29_FINAL.pdf
https://stats.oecd.org/Index.aspx?DataSetCode=EDU_CLASS

https://assets.publishing.service.gov.uk/government/uploads/system/uploads/attachment_data/file/901803/Weekly_COVID19_Surveillance_Report_week_29_FINAL.pdf

https://stats.oecd.org/Index.aspx?DataSetCode=EDU_CLASS

## Acknowledgements

We thank Joshua Weitz and Marm Kilpatrick for helpful critiques of an earlier draft.

